# Core elements of the Positive Health dialogue during patient consultations: a qualitative study exploring expert and user opinions in the Netherlands

**DOI:** 10.1101/2024.07.19.24310586

**Authors:** Eva Aalbers, Miriam de Kleijn, Marja van Vliet, Marieke Spreeuwenberg

**Author notes:** Corresponding author: Eva Aalbers.

## Abstract

**Objectives:** The Alternative Dialogue (PHD), a dialogue technique which is based on Positive Health, is considered a potential method to improve patient-centred care within the Netherlands. This study aims to provide clarity on the core elements of the PHD and aims to assess to what extent the practical application of the PHD aligns with these core elements.

**Design:** An exploratory qualitative study using semi-structured interviews.

**Setting:** The study was conducted among health professionals working in primary and secondary care in the Netherlands.

**Participants:** Thirteen experts were purposely sampled and included based on their involvement in the development, implementation or research of the PHD. Additionally, seven users, who applied the PHD in patient consultations and worked as a primary or secondary health professional participated, both self-selected and purposively sampled. They were included if they participated in a Positive Health training.

**Results:** The analysis revealed consensus among experts and users about prioritizing the patient’s perspective, adopting a holistic health view, and promoting self-management and empowerment as main guiding principles. Consensus was also found regarding professional attitude, goals and outcomes and implementation conditions. Variability was observed in the role of behaviour change support as a guiding principle. Further, the PHD as intended by its developers seems more structured and comprehensive than often applied in practice. Discrepancies also emerged regarding target patient groups and applicable settings, highlighting the need for customization and tailored guidance within diverse contexts.

**Conclusions:** While there is alignment on the main guiding principles of PHD, there are varying opinions regarding its specific tools and techniques. Clarity in terminology and delineation of the PHD, along with customization for diverse contexts, is crucial to address these challenges and to determine its effectiveness. This study provides initial insights to inform future research and practice in PHD implementation.

**STRENGTHS AND LIMITATIONS:**

- As far as known, this is the first scientific study to systematically study the alignment of the core elements of the PHD as regarded by experts, with the practical application across a variety of professions
- The deliberate exclusion of a pre-formulated definition of the PHD in the selection criteria attempted to attain a realistic and impartial reflection of the actual application. However, it resulted in a wide variety of interpretations and, in combination with the amount of participating experts, might have caused an abundance of results and a difficulty in identifying and isolating the PHD as a uniformly implemented intervention.
- The analysis of data was guided by the Framework Methods to systematically guide the process of theme abstraction and data interpretation.
- The selection methods may have resulted in participation bias and/or reporting bias, which may have affected the results.
- Due to the nature of the PHD and the effects the Dutch culture and the Dutch healthcare system have on the application of the PHD, the generalizability of the study results cannot be guaranteed.

## INTRODUCTION

As a response to the illness-oriented and biomedical character of the WHO definition of health, Huber et al. (2011) proposed a new conceptualization of health, in which health is formulated as ‘*the ability to adapt and to self-manage in the face of social, physical, and emotional challenges’* [1]. This dynamic concept was considered a promising approach by the Dutch government to address the challenges of promoting well-being and reducing the burden of (chronic) disease [2]. Therefore, Huber et al. (2016) operationalized this concept into Positive Health (PH). PH regards health as a broad concept, consisting of six dimensions: *bodily functions, mental well-being, meaningfulness, quality of life, participation and daily functioning* [3]. It focuses on well-being and meaningfulness, even in the presence of a chronic condition. According to PH, awareness of one’s health situation, including one’s strengths, weaknesses, preferences, and desires, forms a cornerstone to enhance resilience and self-management [4]. To facilitate practical application, the ‘My Positive Health’ (MPH) Tool was developed, in which the six dimensions are visualized in a spiderweb diagram (see Figure 1) [4,5]. With the online version of the MPH tool, individuals can score 44 statements across the six dimensions using an 11-point Likert scale, after which a ‘health area’ is pictured in the spiderweb diagram. Besides, a paper based version of the tool is available on which each dimension can be rated. The online and paper-based MPH tools are available in adult, adolescent, children’s versions and in a version for people with lower reading skills. In addition, international versions are available in English, German, French, Spanish, Japanese and Icelandic language [6].

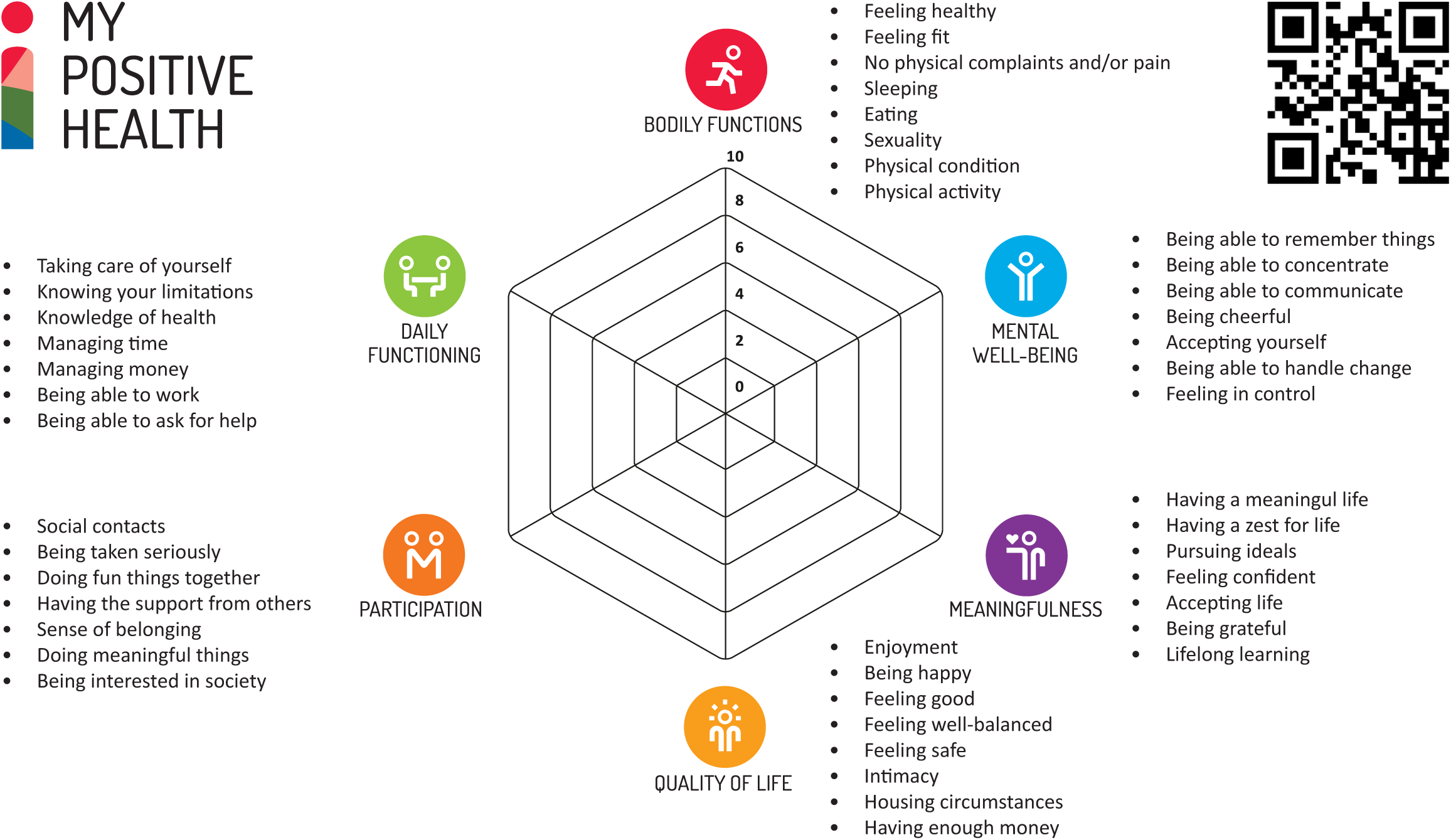

Use of the MPH dialogue tool is part of the so-called Positive Health dialogue (PHD) (or in Dutch: ‘het andere gesprek’). Originally, this PHD as proposed by Huber and colleagues consists of three distinct steps [4]. The initial step is the completion of the MPH tool by the individual to obtain an overall insight in one’s perceived health. The second step is the conversation between an individual and his or her healthcare provider in which the individual’s health needs, desires and values are discussed, along with what is of importance to the individual. In the third step, individuals explore and discuss with their health provider which courses of action to take. In this article, we refer to this application of the PHD as the ‘3-step method’. The PHD is currently applied in primary and secondary care settings, such as by general practitioners, nurse specialists, physical therapists and social workers across the Netherlands [7,8]. Applying the PHD was found to result in a better perceived quality of care, less referrals to the hospital, less prescriptions and a higher job satisfaction [9–11]. Overall, this resulted in lower overall healthcare expenditures [12]. However, there is great diversity in the understanding and application of the PHD, and the role of the MPH tool and 3-step method during the dialogue [8,13,14]. This lack of clarity increases the risk of multi-interpretability and implementation variation, and it remains unclear how this influences the effectiveness of the use of the dialogue tool. According to the study of implementation science, uniform application of core components is crucial for programs and interventions to be effective and sustainable [15,16]. Additionally, as described by Rogers, interventions are often adapted by implementers to better suit their situation and ideas, in a process labelled as ‘reinvention’[17]. Although reinvention could enhance adoption, it could also result in violation of the intervention as intended by the developers, and the effectiveness of the intervention could be impeded if active elements or working mechanisms are compromised[18]. Finally, a recent review study shows that PH and the PHD are regarded as a potential approach to facilitate the shift from an illness-oriented to a health-oriented and person-centred healthcare system within the Netherlands, resulting in a widespread application of PH and the PHD among professionals within health and social care [13]. This is reflected by the high use of the MPH tool, with over 450 000 users of the online tool since its inception in 2016 [5]. Along with this trend, there is a stronger desire to scientifically study the effectiveness of the PHD, for which uniformity in the interpretation and application of the PHD is needed. This all requires a clearer demarcation and identification of the core elements of the PHD, in which the practical application aligns with the developers intention. Core elements are defined as ‘*essential functions or principles, and associated elements and intervention activities that are judged necessary to produce desired outcomes*’ [19]. This study therefore aims to provide clarity on the (core) elements of the PHD and its practical application, by addressing the following research question:

To what extent does the practical application of the PHD correspond with the core elements of the PHD according to experts?

In order to answer this question, the following sub-questions will be addressed:

1. What are the core elements of the PHD according to experts?
2. Which core elements are included in the practical application of the PHD?

The findings of this study will serve to enhance implementation of the PHD, by providing more clarity on the core elements and insight in the congruence of theoretical principles and practical application. Moreover, clarity on the core elements and the uniformity of application can be used as input for further research on treatment fidelity levels and on measuring the effects of applying the PHD.

## METHOD

### Design

This study is an explorative qualitative study, using semi-structured interviews. The study is reported in accordance with the Standards for Reporting Qualitative Research (SRQR) guidelines and checklist [20].

### Setting

This study was conducted in the Netherlands, where the concept of Positive Health by Huber et al. (2016) originated. In 2015, the Dutch Institute for Positive Health (iPH) was established as a non-profit organization dedicated to promoting the dissemination and implementation of PH. Collaborating with training partners, iPH currently offers two training programs designed for professionals within healthcare and social domains: the foundational training (‘Basic module’) to familiarize participants in working with PH and conducting the PHD, and a teacher training (‘Train-the-trainer module’) which aims at training participants to provide the basic training [21]. While the exact number of professionals who have completed the Basic Module training remains unknown, as of July 2024, 320 professionals have successfully completed the train-the-trainer module and are certified to conduct Basic Module training. To increase the comparability of the results and because PH was initially developed within the healthcare sector, this study focuses on primary and secondary care.

### Participants and data collection

Semi-structured interviews were held between July 2021 and June 2022. Participants for the expert interviews were purposely sampled after consultation with iPH. Experts were selected based on their expertise and involvement in the PHD. Inclusion criteria for experts were: (a) involvement in PHD development, or (b) participation in PHD dissemination and/or implementation, or (c) research conducted on PHD. For the user interviews, participants were conveniently recruited using respondent driven sampling. Invitations to participate were distributed via LinkedIn and 1Sociaaldomein (a Dutch digital platform for professionals in health and social care). Additionally, due to an initial low response rate, user participants were actively approached within the network of iPH. Inclusion criteria for the users were: (a) completion of the ‘basic module’ training by iPH, and (b) application of the PHD in patients or client interactions, and (c) working within primary or secondary healthcare practice in the Netherlands. The criteria regarding training were included to ensure familiarity with the concept of PH and the PHD. No predefined definition of the PHD was provided to allow for diverse interpretations. Interviews were held online, in Dutch, via Microsoft Teams or telephone, and were audio-recorded on a separate device. AE conducted the interviews making use of interview guides, which contained main topics and question prompts. The emphasis on different topics was adjusted based on the interviewee’s expertise and experience.

### Participant characteristics

A total of 25 experts were approached via e-mail, of which 15 were willing to participate. The reasons for non-participation were non-responding to invitation (n=2), participant no longer being in function (n=2), unwillingness to participate (n=1) and difficulty in scheduling an appointment (n=5). Two experts were excluded for not meeting the inclusion criteria. 12 semi-structured interviews were held with a total of 13 experts, with an average interview duration of 51:40 minutes (SD=11:30). Additionally, seven interviews were held with users of the PHD, with an average duration of 48:47 minutes (SD=12:40). Participant characteristics can be found in Table 1 and Table 2.

**Table 1.** Participant characteristics part A - Expert interviews.

| Participant | Occupation | Involvement<br>in the topic | First-hand experience<br>with conducting the PHD |
| --- | --- | --- | --- |
| E1 | Former General Practitioner/Researcher | Development | Yes |
| E2 | Researcher and Psychologist | Implementation | No |
| E3 | Strategic advisor | Implementation | No |
| E4 | Advisor Positive Health | Development | No |
| E5 | Program Leader Knowledge Institute | Implementation | Yes |
| E6 | Policy advisor mental health | Implementation | Yes |
| E7 | Program manager prevention | Implementation | No |
| E8 | Researcher | Research | No |
| E9a | Senior researcher | Research | No |
| E9b | Researcher | Research | No |
| E10 | General Practitioner | Development | Yes |
| E11 | Coordinator | Implementation | No |
| E12 | General Practitioner | Development | Yes |

**Table 2.**
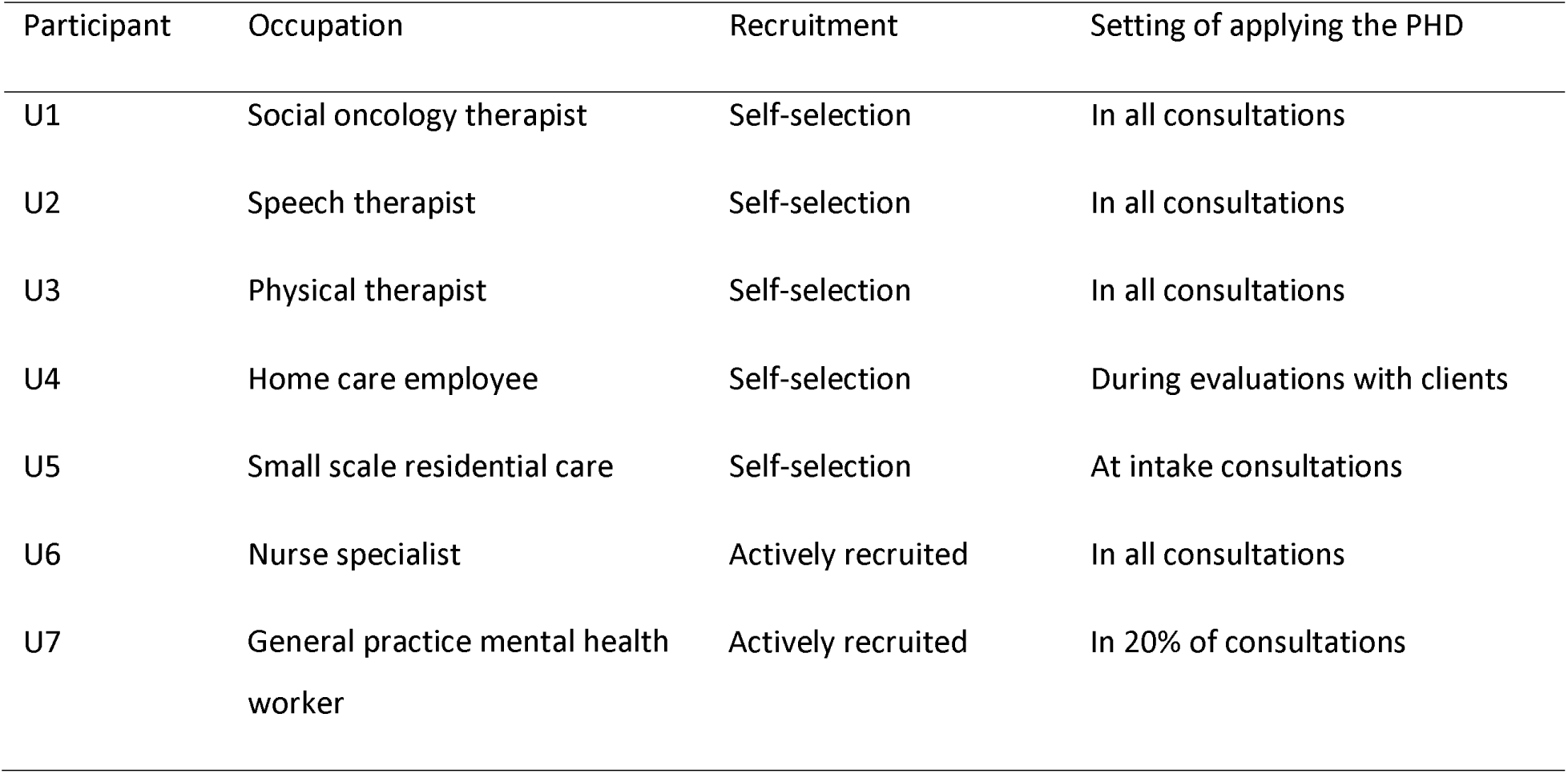
Participant characteristics part B - User interviews.

| Participant | Occupation | Recruitment | Setting of applying the PHD |
| --- | --- | --- | --- |
| U1 | Social oncology therapist | Self-selection | In all consultations |
| U2 | Speech therapist | Self-selection | In all consultations |
| U3 | Physical therapist | Self-selection | In all consultations |
| U4 | Home care employee | Self-selection | During evaluations with clients |
| U5 | Small scale residential care | Self-selection | At intake consultations |
| U6 | Nurse specialist | Actively recruited | In all consultations |
| U7 | General practice mental health worker | Actively recruited | In 20% of consultations |

### Data analysis

The expert and user interviews were conducted consecutively and analysed between May and July 2022 using the framework method [22]. This method offers a structured and transparent process, making it particularly suitable for organizing and interpreting data from multiple sources, such as expert and user interviews. Additionally, the framework method facilitates the identification of key themes and patterns within the data, enabling researchers to derive meaningful insights even when faced with complex or large datasets. Interviews were transcribed manually, and transcripts were anonymized before being uploaded to NVivo for analysis. Data were coded inductively by EA using descriptive codes, which emerged from both the transcripts and observations during the interviews. Themes were extracted from the descriptive codes, which were further categorized into main themes. This was done by an iterative process in which identified main themes and themes were discussed with the research team. Based on the themes, a framework matrix was developed by EA to summarize responses per participant per theme. To enhance credibility, content of the summaries in the framework was peer-reviewed by MS. Subsequently, the framework matrices underwent member-checking by the experts, with six experts providing minor remarks. During data analysis, equal weight was given to all participant responses regardless of their characteristics. The results from both expert and user interviews were combined and reported collectively to improve readability. A peer reviewer was consulted to ensure the reliability of both the framework matrices and the study’s results section.

### Ethical considerations

The research was conducted as a part of the Health Education and Promotion Master program from Maastricht University. Ethical approval was granted by the Research Ethics Committee of Maastricht University Faculty of Health, Medicine and Life sciences, registered under license FHML/HEP_2021.574. Participation was entirely voluntary, and data was handled confidentially. Prior to interviews, participants were briefed on the study’s purpose, privacy and data handling procedures, and provided informed consent.

## RESULTS

Framework analysis of the expert interviews revealed 24 themes - or core elements of the PHD - divided across six main themes: Guiding principles, Professional attitude, Setting and target group, Use of tools and methods, Goals and outcomes, and Implementation conditions. All main themes and themes are detailed below, and a schematic representation can be found in Table 3. Elements marked in italics were found not always to be applied in practice according to the user interviews.

**Table 3.**
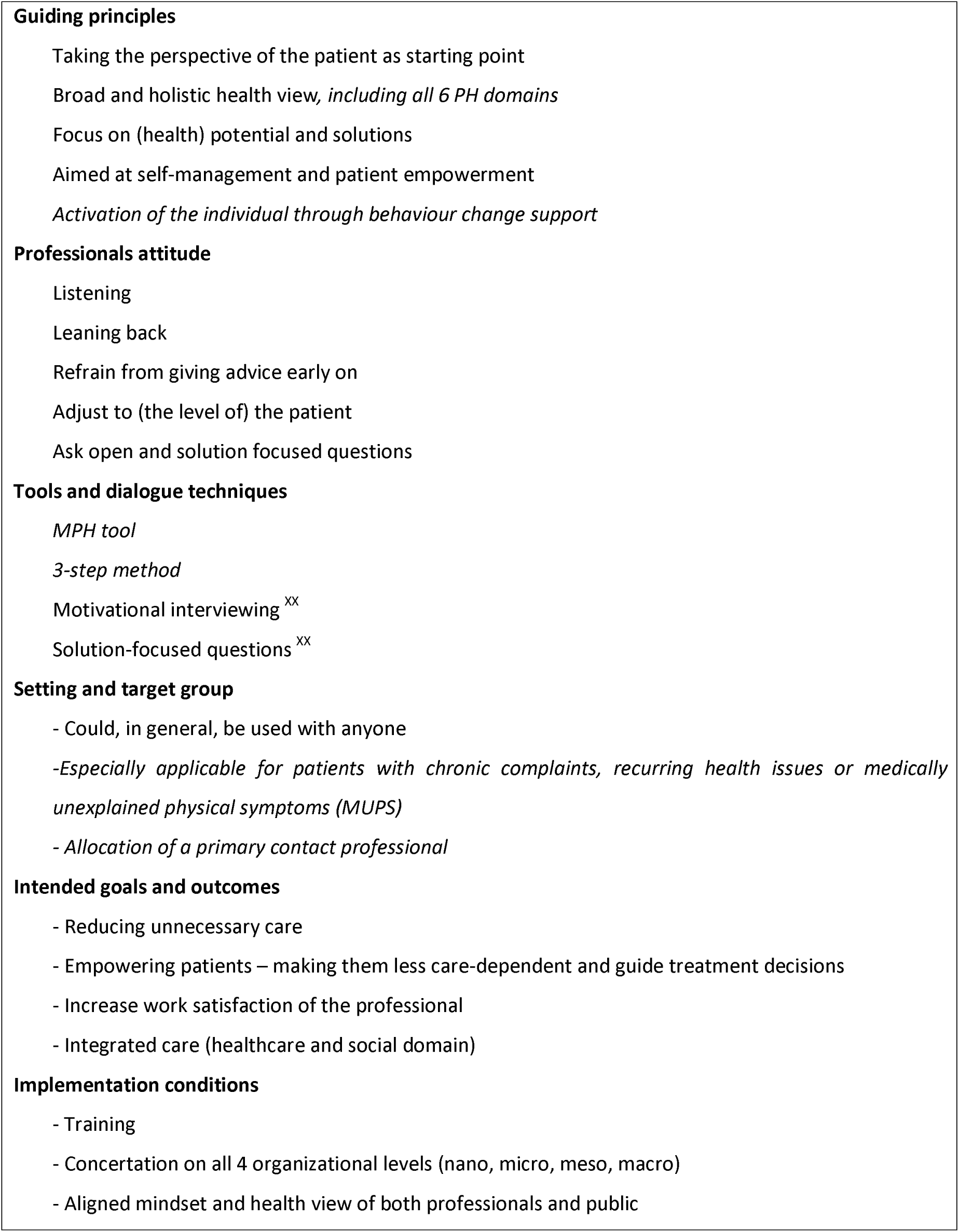
Core elements of the Alternative Dialogue.

### Guiding principles

The expert interviews disclosed the following guiding principles of the PHD: Taking the patient’s perspective (n=13); A broad and holistic view of health (n=11); a focus on solutions and opportunities instead of problems (n=10) and self-management of the individual (n=13).

<u>Taking the patient’s perspective</u> entails centralizing what is important to the individual. It is mostly expressed in the attitude of the professional during the dialogue, which is addressed below. As participant E8 stated: ‘*The alternative dialogue, the core and intention of that is, what I believe, that you really connect to the other person and instead of reasoning from a supply perspective, and from your own professional frame (….) You need to ask people: Who are you actually? What is your situation? What do you need, what are your wishes? What is realistic? What do you perhaps want to change, and how can I support you in that?* Two experts specifically mentioned the importance of first and foremost acknowledging and appropriately addressing emerging medical issues and considering the potential conflict between the patient’s preferences and the medical profession. Centralizing the patient’s perspective was seen as defining characteristic of the PHD by 6 users.

According to 11 experts, <u>the broad and holistic health view</u> incorporates all the six domains of PH in the conversation, which was said to provide both the individual and the healthcare professional (HP) with insight in the individual’s (positive) health and overall situation, as well as how all different domains are connected. Both experts and users recognized that, in practice, not all domains and underlying MPH aspects or statements were considered relevant for every profession. Therefore, not all domains are always included, or are adapted to fit the HPs’ profession. As U2 stated: ‘*I am working on adjusting the spider web questions to the speech therapy field. Because then it would be a tool that is usable in speech therapy, on how the clients experience their logopedic health.’*

According to both experts (n=10) and users (n=6), the focus of the PHD should be on <u>solutions, health potential, and positive change instead of problems and (isolated) health issues</u>. Adding to that, the solutions or proposed action should preferably be coming from the individual instead of the HP.

The focus on <u>self-management</u> was mentioned by all participants. The PHD is intended to assist individuals in identifying various aspects of their health and well-being, thereby increasing their awareness of their own situation and needs. However, experts and users differed in opinion to which extent <u>patient activation</u> is a deliberate part of the PHD. The formulation of concrete, small action steps and the guidance of actual behaviour change was seen as an important element of the PHD by all experts involved in the development of the DA. Nonetheless, only four of the other experts and none of the users alluded to this as an indispensable part of the PHD. Regardless, both users (n=6) and experts (n=10) stated that the PHD should result in patient empowerment, by touching upon the intrinsic motivation and the individuals’ locus of control.

### Professionals’ attitude

Both experts (n=13) and users (n=5) viewed the attitude of the HPs and the use of conversational techniques as key features of the PHD and its application. According to the experts, the HP should <u>mostly listen</u> (n=5), <u>sit back</u> (n=7), <u>refrain from giving advice early on</u> (n=10), <u>adjust to (the level of) the patient</u> (n=8), and <u>ask open and</u> <u>solution focused questions</u> (n=4). *‘It requires sitting on one’s hands. It requires allowing awkward silences, and facilitating instead of steering the conversation’* (E11). Five experts stated that it is a skill anyone could learn, although mastering the PHD requires extensive training, experience and regular (self-)reflection. Most experts (n=10) agreed that working with the PHD requires a significant shift in attitude and mindset of the HP, and the HP should be open to the holistic and broad health perspective, and to the importance of patients’ self-management. This was also reflected in the user interviews. As U6 stated: ‘*Positive Health is not some kind of trend, something that needs to be done once and then it’s over with. It is something that should be in your heart and soul, it should land in your DNA.’*

### Tools and dialogue techniques

Opinions on the use of <u>the MPH tool</u> differed among participants. Some (n=6) experts identified the MPH tool as a defining component: ‘C*arrying out the alternative dialogue is having a dialogue based on the results of the MPH tool’* (E11), whereas 2 experts specifically opposed that: *‘It is not about using the MPH tool to be applying positive health, but it is about carrying out what it stands for’* (E05). Three experts specifically mentioned that the focus of the conversation should not be on the lowest scores on the MPH tool, but rather on what individuals wanted to talk about. Three users regularly used the complete, 44 question version of the MPH tool. All users expressed that they keep the six PH domains in the back of their mind during the consultations, but some expressed that the completion of the entire MPH tool during consultations is not always doable or appropriate. All but one user expressed that they either adapt the MPH tool to their setting, or would prefer a more setting-specific tool, since not all domains or statements are in their opinion applicable to their setting. Opinions also differed regarding the <u>3-step method</u> of the PHD, encompassing a reflection on overall health by MPH tool, identification of preferences, and selection of courses of action. Although the experts propose a structured dialogue, most users (n=5) expressed that in practice, the dialogue requires adaptation to the individual and the circumstances. Three expert interviews revealed that, even though the PHD is originally based on the use of the 3-step model, in practice the term ‘Alternative Dialogue’ is regularly used to describe any consultation in which a patient-centred, broad view is applied. Two experts explicitly noted that we should be hesitant to call any dialogue based on the MPH tool the PHD. However, there was no consensus regarding the possibility and desirability to demarcate the PHD. Six experts stated that the formulation of guidelines or a type of framework is possible, while at the same time recognising that the flexibility and adaptability is seen as a strength of the concept. As expert 9a stated: *‘I don’t know if you should really delineate it for what it should be. On the one hand I would say yes, because that makes it measurable, if everyone does the same, (….) On the other hand, I think the Alternative Dialogue is also about being able to have your own interpretation of it, as a HP, together with the clients.’* From the users perspective, flexibility and adaptability to the setting and/or target group were highly valued, which was reflected in the variety of applications and adaptations.

Key dialogue techniques included in the PHD, as highlighted by the participants, were motivational interviewing[23] (experts n=3; users n=4) and use of solution-focused questions[24] (experts n=4; users n=5).

### Setting and target group

The setting in which the PHD is and could be applied, was found to depend on how the PHD is defined. Although the PHD was said to be <u>suitable for everyone</u>, especially experts who defined the PHD based on the completion of the MPH tool and the 3-step method, <u>mentioned a specific target group</u> with whom the PHD could be particularly useful (n=7). For example patients with medically unexplained physical symptoms and/or patients for whom the biomedical model will not offer a solution. As participant E14 shared: ‘*Out of the 170 people I used it with, one third were people with recurring symptoms or complaints such as headaches, back pain, sleeping problems, dizziness etc. One third had mental health problems, and about one third either had a chronic condition or the conversation focused on lifestyle changes. And another 10% was for introductory conversations with new patients.’* Three experts stated that it doesn’t matter who applies the PHD, as long as someone in the healthcare chain does it. One expert stated that all HPs should apply some form of the PHD, while another expert specifically opposed the idea of the PHD being implemented in all consultations. Five experts suggested the <u>allocation of a central key person</u> who retains coordination, to prevent repetition and protect patients’ privacy, especially when applying the comprehensive 3-step model PHD. There was no consensus among both experts and users on whose responsibility this should be, since many professions have limited time resources, and extensive coaching might not be the HP’s expertise or preference. In practice, all but one user stated to apply the principles and mindset of the PHD with all their patients in all consultations.

### Intended goals and outcomes

The interviews harvested a wide variety of intended outputs, outcomes and goals, which were related to the setting in which the PHD was applied. Firstly, eight experts mentioned the goals of <u>reducing unnecessary care</u> by assisting patients to first explore what they can do themselves to improve their health and resilience, by better matching the needs and wishes of the patient and by uncovering actual underlying or contributive issues. Secondly, 10 experts mentioned <u>empowering patients</u> as a goal of the PHD, which helps to make them less care-dependent (n=10) and supports them to make a treatment decision according to the needs and wishes of the patients (n=4). Thirdly, six experts mentioned <u>increased work satisfaction for HPs</u> as an outcome. Three experts recognized the need for concrete actions and follow-up plans, whereas one other expert stated that there is not always a concrete output, since this is subject to the course of the conversation and the information that is brought up by the individual. Additionally, experts mentioned <u>integration of healthcare with the social domain</u> (n=5) as potential output. The users’ goals and outcomes mainly aligned with the expert views. The users mainly applied the PHD in order to better match their provided care to the perspective of the patient. One user added that the effects of the PHD are not always measurable (e.g. in the case of refraining from treatment) and/or are not always directly and solely attributable to the PHD. As U6 stated: *‘It is about that, when people leave, that you see that they are stronger, they stand up straight, that they don’t leave feeling disheartened. That they feel energetic. These are all very subjective, I don’t think there is anything concrete…. What would then be the outcome measure?’*

### Implementation conditions

Experts mentioned prerequisites of implementing the PHD, namely: training (n=11), organizational concertation (n=11), and a collective, societal change in the way health and healthcare are regarded (n=8). All experts saw <u>adequate training</u> of the HP as an important prerequisite. As participant 1 stated: ‘*Having working experience, and, in general, life experience, makes it easier, but being trained is still essential.’* Focus during the training should be on understanding the concept of positive health, practicing the conversational techniques, receiving feedback on it and continuous self-reflection or peer-to-peer support. Additionally, experts mentioned there should also be attention to the overall attitude of the HP and how to place themselves and the PHD within their organization and work context. Four experts advocated for the incorporation of the training elements into regular HPs’ education. Users expressed that the training was valuable for practicing the PHD, while four users also expressed a need for a more in depth, tailor-made and context-specific training and implementation support.

Ten experts stated, directly or indirectly, that <u>organizational coherence and co-operation</u> is needed on several levels. Where discussed, this view was supported by the users. Besides the use of the PHD in the consultation room, there should be an aligned culture and awareness in HP practices or organizations. Further, mutual awareness, interprofessional communication and cooperation, and an adequate referral system within the neighbourhood and district are considered important by the experts. At last, regional and national organizations need to support an integrated care approach. Two users expressed struggles to match the execution of the PHD with administrative requirements and a lack of financial compensation from healthcare insurance companies. Both users and experts were divided on whether the PHD requires more time than usual consultations. Although admittedly, the concept of the PHD was not regarded as new or groundbreaking by four users and four experts, Positive Health was described as bridging the gap between the medical and social domain, and having a common, shared language was said to enable better co-operation on all above mentioned levels.

Additionally, five experts and two users noted that both the HPs as well as the general public need a <u>change in</u> <u>their view of health and healthcare</u>: ‘*The way how we, as HPs, have been dealing with diseases, health behaviour and clients and patient for years, for centuries, has also led to patients and clients who are not used to thinking about what they actually want, what is important to them’* (E5) *‘Because they go the doctor with an old biomedical frame of mind. They tell their problem, and they expect a solution from the doctor’*(E14).

## DISCUSSION

The aim of this study was to provide clarity on the core elements of the PHD based on expert interviews, and to what extent these are applied in practice according to its users. Based on findings from both expert and user interviews, several shared core elements were identified. Experts and users agreed on key guiding principles, including prioritizing the patient’s perspective, adopting a broad health view, focusing on potential and solutions, and promoting self-management and empowerment. Agreement was also observed regarding the attitude of the HP (listening, leaning back, refraining from giving advice, adjust to the patient, and ask open and solution focused questions), intended goals and outcomes (reducing unnecessary care, empowering patients, increase HP work satisfaction, and fostering integrated care), and implementation conditions (training, concertation on all organizational levels, and an aligned mindset and health view of both the professional and the public). However, variability was observed in the interpretation of the tools and dialogue techniques of the PHD, especially regarding the flexibility in the use of the MPH tool and the 3-step method, both among experts and users. This suggests potential ambiguity in application of the PHD. Furthermore, discrepancies were identified between developers’ intentions and practical application regarding the inclusion of patient activation through behaviour change guidance. Additionally, while experts emphasized that the PHD was especially useful for specific target groups, such as patients with chronic complaints or medically unexplained physical symptoms, users did not reach a consensus on this matter. Furthermore, there was disagreement within and between experts and users regarding who conducts the PHD conversation and the allocation of a primary contact person.

These findings lead to the following reflections and recommendations:

Firstly, according to both experts and users, there is a strong emphasis on the conversational techniques and overall attitude and mindset of the HP during the PHD. This finding to some extent matches research on patient-centred care, where a study by Kitson et al [25] identified personal qualities such as ‘*being polite’, ‘good etiquette’, ‘good manners’, ‘being respectful’, ‘sensitive’, ‘welcoming’* as part of one of the core elements of patient-centred care. This finding, in combination with the participants’ input, emphasizes the need for thorough training, as research suggests that these conversational techniques are trainable skills [26–29]. In Miller et al. (2001) experts indicated motivational interviewing and the use of solution focused questions as applicable methods, and most users stated to apply these. However, observations of consultations would be recommended to analyse more thoroughly to what extent these conversational techniques, in combination with a patient-centred attitude and mindset, are actually applied and with what level of fidelity, since HPs may not accurately report reality [26].

Secondly, the Dutch term for the PHD - ‘het andere gesprek’ - seems to be interchangeable for both a structured, intervention-like application of the PHD based on the MPH tool and the 3-step method as intended by the developers, as well as for the overall concept of working with a ‘Positive Health mindset’. This ambiguous use of terms might create confusion amongst parties. Yaron et all (2021) stated that ambiguous linguistics contributed to multiple interpretations of the concept of Positive Health [14]. This lack of clarity complicates efforts to implement Positive Health practices effectively. Miller and Rollnick [18] stated that, for Motivational Interviewing, confusion about the terminology and constitution impeded the quality of scientific research, clinical practice and training. Given the general use of the term ‘het andere gesprek’ as a more general mindset, it is advisable to refine or reconsider the terminology for the more comprehensive dialogue, which is based on the 3-step method. Simultaneously, clarity is needed on what qualifies as PHD within Positive Health (PH), similar to how guidelines were established for Motivational Interviewing [18]. This study and the Handbook Positive Health can thereby serve as a basis [4]. Furthermore, if the comprehensive version of PHD would be considered unsuitable for every practical context and setting, we recommend reassessing the feasibility and effectiveness of adopting a less time-intensive approach without diminishing the core elements and underlying working mechanisms. Subsequently, careful consideration should be given to determining where the responsibility for conducting the PHD lies, and how mutual cooperation and information exchange can be effectively organized.

Thirdly, this study revealed a lack of consensus regarding target patient groups and applicable settings of the PHD. Respondents in our study indicated that users perceive flexibility to adapt the PHD, including the MPH tool itself, to suit their particular setting and profession. This variation in application may lead to a wide range of approaches. At the same time a need was expressed by the user respondents for a clearer scope and delineation of the MPH tool in clinical practice. This was also found in a study by Bock et al. (2021) in which medical specialists expressed uncertainties regarding appropriate target groups and settings for implementing PHD, as well as the most effective utilization of the MPH tool [7]. A potential explanation for this lack of clarity could be that the main developers of the PHD have a background as General Practitioner, while all participating users represent a wide range of professions. Although the experts provided a general outline of the PHD, they also acknowledged the necessity for (further) customization of the MPH tool and the PHD for use within different settings, as well as the need for support in this process. As expressed by Yaron et al. (2021), ‘Tailoring is crucial in the application of Positive Health, due to the unique character and context of each organization.’ [14] This becomes increasingly important as PH is being adopted across various domains, not only within healthcare but also beyond. This includes the social domain, work, school, and the living environment [13]. Therefore, further research is needed to identify if, how and to what extent the PHD and MPH tool should and could be tailored to each specific setting. Research is needed among both end-users (patients and citizens) and their caregivers to explore in which extent the need for tailor-made versions is expressed among each population. Engaging end-users and relevant professionals from different domains during this research will enhance the methods’ suitability and accessibility.

Fourthly, this study provides a first draft of the core elements of the PHD. The working mechanisms and effects of (the combination of) these elements are yet to be discovered, for example through practical assessments, empirical validation, and by formulating a more in depth, literature-based theory of change. This also requires a clear formulation of the PHD, including measurable goals and preferred outcome(s) to determine to what extent the proposed working mechanism actually fulfils its predicted outcomes. One of the theories underlying the concept of the PHD, that could act as a theory of change, is the Sense of Coherence [6,30,31]. By not following the 3-step method, it is possible that not all three aspects of the Sense of Coherence (comprehensibility, manageability and meaningfulness) are covered. Building on the findings of this study, a more elaborate checklist or skill code to assess application fidelity could be developed, as was done for motivational interviewing [32,33].

Fifth, this study found that behaviour change support and patient activation are not always considered part of the PHD. Therefore, further research is warranted if these aspects are one of the core elements of the PHD and thus should be part of PHD training. Additionally, recent work by Sponselee et al. (2024) underscores the need to take specific skills and capacities into account, such as executive functioning, non-cognitive skills, and health literacy. These abilities play a pivotal role in empowering individuals to effectively manage their health conditions. Recognizing the variability in individual capacities, it seems necessary to provide tailored guidance and support during the PHD[34].

Finally, this study found that, for a successful application of the PHD, organizational and societal changes in culture are considered important, and the PHD should not be approached as a stand-alone intervention. This is in line with dissemination and implementation theories such as the Consolidated Framework for Implementation Research [32] and the Fleuren model [35] which, both in their own words, state that the effectiveness and implementation rates of intervention are not only dependent on the characteristics of the intervention, but are also affected by characteristics of the inner setting, the outer setting and the individuals. The need for good cooperation, referral options and societal alignment were also found by other researchers [13,14].

## STRENGTHS AND LIMITATIONS

As far as known, this is the first scientific study to systematically study the alignment of the core elements of the PHD as regarded by experts, with the practical application across a variety of professions. This study included participants with varied backgrounds and perspectives. Although the number of user participants was limited, five of the expert participants had personal experience with the application as well, and thereby indirectly more user experiences were included. To allow for different perspectives, both experts related and not related to IPH (who provides training on PHD) were included in the study. The deliberate exclusion of a pre-formulated definition of the PHD in the selection criteria might have resulted in a wide variety of interpretations and might have caused an abundance of results, although this was done in an effort to reflect the real-world situation. The length of the research period might have affected the discrepancy between expert and user results, since most of the expert interviews were conducted about 8 months before the user interviews. Further, the study made use of interviews instead of observations, which might have caused reporting bias. While there is a risk for participation bias, due to self-selection of the user participants, the variety in interpretation and application and the alignment of the results with previous research suggests a sufficient representation. To improve credibility, member checks and peer reviews were conducted.

## CONCLUSIONS

In conclusion, the study aimed to explore the extent to which the practical application of the PHD aligns with its core elements as identified by experts. The findings underscored a convergence of guiding principles, professional attitudes, intended goals, and implementation conditions between experts and users. Notably, both groups emphasized the importance of prioritizing the patient’s perspective, adopting a holistic health view, and promoting self-management and empowerment as guiding principles. However, variability surfaced in the interpretation and application of specific tools and techniques, particularly regarding the flexibility in using the MPH tool and the 3-step method. The PHD as proposed by the developers seems more structured, and more comprehensive compared to the way the PHD is applied in practice. This ambiguity highlights the need for clarity in terminology and delineation of the PHD, especially as its application expands. Moreover, discrepancies were evident concerning target patient groups and applicable settings, suggesting the necessity for customization and tailored guidance within diverse contexts. This study initials insights to address these issues.

## Author contributions

MS drafted up the study. EA collected and analysed data under supervision of MdK and MS. EA and MvV wrote the manuscript in collaboration with all co-authors. All authors commented and agreed on the final version.

## Data sharing statement

Reasonable requests for sharing data can be made by sending an email to the corresponding author.

## Patient and Public involvement statement

Patients and/ or the public were not involved in the design, conduct, and reporting of the study.

## Data Availability

All data produced in the present study are available upon reasonable request to the authors

## REFERENCES

1. Huber M, Knottnerus JA, Green L, et al. How should we define health? Bmj 2011;343:d4163. doi: 10.1136/bmj.d4163 [published Online First: 2011/07/28]

2. Johansen F, Loorbach D, Stoopendaal A. Exploring a transition in Dutch healthcare. J Health Organ Manag. 2018 Oct 8;32(7):875–90.

3. Huber M, van Vliet M, Giezenberg M, et al. Towards a ‘patient-centred’ operationalisation of the new dynamic concept of health: a mixed methods study. BMJ Open 2016;6(1):e010091. doi: 10.1136/bmjopen-2015-010091 [published Online First: 2016/01/14]

4. Huber MAS, Jung HP, Van den Brekel-Dijkstra. Handbook Positive Health in Primary Care: The Dutch Example. Houten, the Netherlands: Bohn Stafleu van Loghum; 2022.

5. https://vragenlijsten.mijnpositievegezondheid.nl/

6. www.positivehealth-international.com/dialogue-tools/

7. Bock LA, Noben CYG, Yaron G, et al. Positive Health dialogue tool and value-based healthcare: a qualitative exploratory study during residents’ outpatient consultations. BMJ Open 2021;11(11):e052688. doi: 10.1136/bmjopen-2021-052688

8. Rijksinstituut voor Volksgezondheid en Milieu (RIVM). Het gebruik van brede gezondheidsconcepten: inspirerend en uitdagend voor de praktijk, 2020.

9. Jung HP, Jung T, Liebrand S, Huber M, Stupar-Rutenfrans S, Wensing M. Meer tijd voor patiënten, minder verwijzingen. Huisarts Wet. 2018;61(3):39–41.

10. Jung HPD, Liebrand SM, van Asten Cd. Uitkomsten van het hanteren van Positieve Gezondheid in de praktijk. Bijblijven 2019;35(8):26–35. doi: 10.1007/s12414-019-0075-x

11. Lemmen CHC, Yaron G, Gifford R, Spreeuwenberg MD. Positive Health and the happy professional: a qualitative case study. BMC Fam Pract. 2021 Dec 1;22(1).

12. Berden C, Kuyterink M, Mikkers M. Beyond the Clock: Exploring the Causal Relationship between General Practitioner Time Allocation and Hospital Referrals [Internet]. Tilburg; 2023 Aug [cited 2023 Sep 20]. (2023). Report No.: 2023–019. Available from: https://research.tilburguniversity.edu/en/publications/beyond-the-clock-exploring-the-causal-relationship-between-genera

13. van Vliet M, de Kleijn M, van den Brekel-Dijkstra K, Huijts T, van Hogen-Koster S, Jung HP, Huber M. Rapid Review on the Concept of Positive Health and Its Implementation in Practice. Healthcare. 2024; 12(6):671. 10.3390/healthcare12060671

14. Yaron G, Spreeuwenberg M, Ruwaard D. Praktijkhandreiking werken met Positieve Gezondheid: Lessen uit Limburg. Utrecht, 2021.

15. Moir T. Why Is Implementation Science Important for Intervention Design and Evaluation Within Educational Settings? Frontiers in Education 2018;3 doi: 10.3389/feduc.2018.00061

16. Eccles MP, Mittman BS. Welcome to Implementation Science. Implementation Science 2006;1(1):1. doi: 10.1186/1748-5908-1-1

17. Rogers EM. Diffusion of Innovations. New York: Free Press 2003.

18. Miller WR, Rollnick S. Ten Things that Motivational Interviewing Is Not. Behavioural and Cognitive Psychotherapy 2009;37(2):129–40. doi: 10.1017/S1352465809005128 [published Online First: 2009/02/06]

19. Blase K, Fixsen D. Core Intervention Components: Identifying and Operationalizing What Makes Programs Work. Washington, DC 20201: US Department of Health and Human Services, 2013.

20. O’Brien BC HI, Beckman TJ, Reed DA, Cook DA.. Standards for reporting qualitative research: a synthesis of recommendations. Academic Medicine 2014;89 doi: 10.1097/ACM.0000000000000388

21. Institute for Positive Health. Gecertificeerde trainers n.d. [Available from: https://www.iph.nl/meedoen/trainingen/gecertificeerde-trainers/ accessed 10-07-2022.

22. Gale NK, Heath G, Cameron E, et al. Using the framework method for the analysis of qualitative data in multi-disciplinary health research. BMC Medical Research Methodology 2013;13(1):117. doi: 10.1186/1471-2288-13-117

23. Miller, W. R., & Rollnick, S. (2012). Motivational Interviewing: Helping People Change (3rd ed.). Guilford Press.

24. de Shazer S & Dolan Y (2007). More Than Miracles: The State of the Art of Solution-Focused Brief Therapy. Routledge.

25. Kitson A, Marshall A, Bassett K, et al. What are the core elements of patient-centred care? A narrative review and synthesis of the literature from health policy, medicine and nursing. Journal of Advanced Nursing 2013;69(1):4–15. doi: 10.1111/j.1365-2648.2012.06064.x

26. Miller WR, Mount KA. A small study of training in motivational interviewing: does one workshop change clinician and clinet behaviour? Behavioural and Cognitive Psychotherapy 2001;29(4):457–71. doi: 10.1017/S1352465801004064 [published Online First: 2001/10/23]

27. Tidhar M, Benbassat J. Teaching Shared Decision Making to Undergraduate Medical Students. Rambam Maimonides Med J 2021;12(4):e0032. doi: 10.5041/RMMJ.10453

28. Tongue J, Epps H, Forese L. Communication skills for patient-centred care: Research-based, easily learned techniques for medical interviews that benefit orthopaedic surgeons and their patients. The Journal of Bone and Joint Surgery 2005;87A:652–58. doi: 10.2106/00004623-200503000-00027

29. Tran B. Strategies for effective patient care: Integrating quality communication with the patient-centered approach. Social and Personality Psychology Compass 2020;15 doi: 10.1111/spc3.12574

30. Lindström B, Eriksson M. Salutogenesis. Journal of Epidemiology and Community Health 2005;59(6):440. doi: 10.1136/jech.2005.034777

31. Nelissen J, Degryse JM. Salutogenese. Huisarts Nu 2015;44:26–31. doi: 10.1007/s40954-015-0012-x

32. Damschroder LJ, Aron DC, Keith RE, et al. Fostering implementation of health services research findings into practice: a consolidated framework for advancing implementation science. Implementation Science 2009;4(1):50. doi: 10.1186/1748-5908-4-50

33. Venner K, Jamieson L. Assessing fidelity in Motivational Interviewing interventions; an overview. 2014

34. Sponselee HCS, Ter Beek L, Renders CM, Kroeze W, Fransen MP, van Asselt KM, et al. Letting people flourish: defining and suggesting skills for maintaining and improving positive health. Front Public Health. 2023;11:1224470.

35. Fleuren M, Wiefferink K, Paulussen T. Determinants of innovation within health care organizations: literature review and Delphi study. Int J Qual Health Care 2004;16(2):107–23. doi: 10.1093/intqhc/mzh030 [published Online First: 2004/03/31]

